# Association of childhood cardiometabolic risk factors with arterial health in adolescence: an 8-year follow-up study

**DOI:** 10.64898/2026.05.08.26352777

**Authors:** Mika Jormanainen, Tina Salmi, Anna Viitasalo, Satu Pekkala, Eija K. Laakkonen, Mustafa Atalay, Tomi Laitinen, Eero A. Haapala, Timo A. Lakka

## Abstract

**Background:** Predictors of arterial health impairment from childhood to adolescence remain largely unknown. We investigated associations of childhood cardiometabolic risk factors with several measures of arterial health in adolescence.

**Methods:** Altogether, 222 children were examined at age 7–9 years and eight years later at age 15-17 years. Body fat percentage (BF%), glucose, insulin, lipids, blood pressure (BP), and inflammation biomarkers were measured and homeostatic model assessment for insulin resistance (HOMA-IR), a metabolic syndrome score (MetSscore), and an inflammation score were calculated at baseline. Pulse wave velocity (PWV) and cardio-ankle vascular index (CAVI) were assessed using impedance cardiography and carotid intima-media thickness (cIMT), carotid distensibility (cDIST), Young’s elastic modulus (YEM), and stiffness index (SI) using ultrasonography. Associations of childhood cardiometabolic risk factors with measures of arterial health were analyzed using linear regression models adjusted for childhood age and sex.

**Results:** BF% was positively associated with PWV (standardized regression coefficient β=0.207, p=0.008), CAVI (β=0.171, p=0.031), cIMT (β=0.146, p=0.034), and YEM (β=0.164, p=0.016). HOMA-IR was positively associated with PWV (β=0.242, p=0.001) and CAVI (β=0.216, p=0.004) and inversely with cDIST (β=-0.162, p=0.015). MetSscore was positively associated with PWV (β=0.266, p<0.001), CAVI (β=0.219, p=0.004), and YEM (β=0.141, p=0.032) and inversely with cDIST (β=-0.140, p=0.035). SBP was positively associated with PWV (β=0.257, p<0.001) and YEM (β=0.156, p=0.018) and inversely with cDIST (β=0.169, p=0.012).

**Conclusion:** Increased adiposity, insulin resistance, elevated SBP, and cardiometabolic risk factor clustering in childhood association of arterial stiffness and reduced arterial distensibility in adolescence, emphasizing prevention of cardiovascular diseases since childhood.

## Introduction

Atherosclerotic cardiovascular diseases (ASCVDs) remain the leading cause of premature morbidity and mortality globally.^1^ These diseases develop through complex, partly interconnected pathophysiological mechanisms that result in increased arterial stiffness, arterial wall thickening, and atherosclerotic lesions.^2^ While the clinical manifestations of ASCVDs, including hypertension, coronary artery disease, ischemic stroke, and heart failure, typically emerge in adulthood after decades of progression, the underlying pathological processes begin already in childhood.^3,4^ Therefore, identifying risk factors for early atherosclerotic changes improves the understanding of pathophysiological mechanisms behind atherosclerosis and helps enhance the identification of individuals at increased risk and develop more effective lifestyle and medical interventions to prevent and treat ASCVDs.^4,5^

One of the most prominent early-life risk factors for ASCVDs is the clustering of cardiometabolic risk factors, often called the metabolic syndrome (MetS).^4^ While MetS is rare in youth with normal body weight, it affects up to 32% of children and adolescents living with overweight or obesity.^5–7^ Key features of MetS include abdominal adiposity, insulin resistance (IR), dyslipidemia, and elevated blood pressure (BP),^5^ along with features like systemic low-grade inflammation and dysregulated adipokine signaling.^6^ Single cardiometabolic risk factors and MetS have been associated with increased risk of type 2 diabetes, preclinical atherosclerosis, ASCVDs and premature mortality in adults.^8–10^ Obesity, MetS, IR, and elevated BP have been associated with increased arterial stiffness in children and adolescents, but evidence remains mixed and is based on cross-sectional studies^3,4,10^ or longitudinal studies with 4-8 years of follow-up.^5,9^ Childhood and adolescence obesity has also been associated with increased carotid intima-media thickness (cIMT) in longitudinal studies, and this association has been partly mediated by IR, increased fasting glucose, and dyslipidemia.^11,12^

While MetS is recognized as a childhood risk factor for poorer arterial health, longitudinal evidence linking individual childhood cardiometabolic risk factors and MetS to measures of arterial health in adolescence remains limited. Therefore, we examined the longitudinal associations of individual cardiometabolic risk factors and MetS assessed in childhood with several measures of adolescent arterial health, including arterial pulse wave velocity (PWV), cardio-ankle vascular index (CAVI), cIMT, carotid distensibility (cDIST), Young’s elastic modulus (YEM), and arterial stiffness index (SI).

## Materials and methods

### Study design and participants

The present longitudinal analyses are based on data on cardiometabolic risk factors and MetS collected from children participating in the baseline examinations of the Physical Activity and Nutrition in Children (PANIC) study and data on the measures of arterial health collected from the same participants attending the 8-year follow-up examinations of the study.^13^

We invited 736 children 7-9 years of age who started the first grade in 16 primary schools of the city of Kuopio, Eastern Finland in 2007-2009 to participate in the study. Altogether 512 children (248 girls, 264 boys) aged 7-9 years, who accounted for 70% of those invited, accepted the invitation and participated in the baseline examinations in 2007-2009. The participants did not differ in sex, age, body height-standard deviation score (body height-SDS), or body mass index-standard deviation score (BMI-SDS) from all children who started the first grade in the city of Kuopio in 2007-2009 based on data from the standard school health examinations performed for all Finnish children before the first grade. We excluded six children from the study at baseline because of physical disabilities that could hamper participation in the intervention or no time or motivation to attend in the study. We also excluded the data of two children whose parents withdrew their permission to use these data in the study. After these exclusions, the baseline study population included 504 children aged 7-9 years, 277 (55%) of whom also attended the 8-year follow-up examinations in adolescence at the age of 15-17 years.

For the present analyses, we excluded participants with conditions potentially affecting cardiometabolic or arterial health, including one child with Klinefelter’s syndrome but none with type 1 or 2 diabetes, and those who had entered clinical puberty by baseline as defined by Tanner staging.^14^ Complete data for the analyses on the associations of individual cardiometabolic risk factors and MetS at baseline with PWV and CAVI at the 8-year follow-up were available for 175 participants (85 girls, 90 boys) and on the associations of individual cardiometabolic risk factors and MetS at baseline with cIMT, cDIST, YEM, and SI at the 8-year follow-up were available for 222 participants (105 girls, 117 boys).

The Research Ethics Committee of the Hospital District of Northern Savo approved the study protocol in 2006 (Statement 69/2006) and its amendment in 2015 (Statement 422/2015). The parents or caregivers of the children gave their written informed consent, and the children provided their assent to participation. The PANIC study has been carried out in accordance with the principles of the Declaration of Helsinki as revised in 2008.

### Assessment of adolescence arterial health

PWV and CAVI were measured at the 8-year follow-up using the CircMon^®^ B202 impedance cardiography device (JR Medical Ltd, Saku Vald, 419 Estonia).^15^ The participants were asked to rest in a supine position for 15 min before the measurement. Next, current electrodes were placed on the distal parts of the extremities, slightly proximal to the wrists and the ankles. Voltage electrodes were placed proximal to the current electrodes, with a distance of 5 cm between the centers of the electrodes. The distal impedance plethysmography was recorded from a popliteal artery at the knee joint level.^15^ The active electrode was placed on the lateral side of the knee joint, and the reference electrode on the calf. The distance between the electrodes was about 20 cm. Alternating electrical current was applied to current electrodes and change in whole-body impedance was measured from voltage electrodes. The CircMon^®^ B202 software estimates the foot of the impedance cardiography signal that coincides with pulse transmission in the aortic arch and the foot of the impedance plethysmography signal that coincides with pulse transmission in the popliteal artery.^15,16^ The CircMon^®^ B202 software calculated PWV (m/s) by dividing the assessed distance (L) by utilizing the measured pulse transit time (Δt).^15^ PWV refers to the speed at which the BP wave travels through the arteries and is a key indicator of arterial stiffness. Higher PWV reflects stiffer arteries and has been associated with an elevated risk of ASCVDs.^17^ CAVI is derived from the arterial stiffness parameter β and is calculated as (2ρ/ΔP) × ln(SBP/DBP) × PWV², where ρ is the estimated blood density, SBP is systolic blood pressure (mmHg), DBP is diastolic blood pressure (mmHg), ΔP is pulse pressure (SBP–DBP), and PWV is heart–ankle pulse wave velocity assessed from the origin of the aorta to the ankle through the femoral artery.^18^ CAVI is calculated as a{(2ρ/ΔP) × ln(SBP/DBP) × PWV²} + b, where ρ, SBP, DBP, ΔP, and PWV are as explained above, and a and b are constants to convert a CAVI value to the value of Hasegawa’s pulse wave velocity, which is compensated for at 80 mmHg of DBP.^18,19^ CAVI has been developed to represent the extent of arteriosclerosis throughout the aorta, femoral artery and tibial artery independent of blood pressure.

Carotid imaging was performed at the 8-year follow-up using the Sequoia^®^ 512 ultrasound scanner (Acuson, Mountain View, CA, USA) with a 14.0 MHz linear array transducer. cIMT and cDIST were assessed by two trained sonographers using a standardized protocol. An electrocardiographic signal was drawn on the ultrasound image by the ultrasound scanner. A 5-second cine loop, which included the beginning of the left carotid artery bifurcation and the left common carotid artery, was recorded and stored for subsequent off-line analyses.^11^ For the assessment of cIMT, the best quality end-diastolic frames, incident with the R-wave on a continuously recorded electrocardiogram, were selected from the video clip. A minimum of three measurements were taken from the far wall of the left common carotid artery approximately 10 mm proximal to the carotid bifurcation to derive the maximal cIMT.^11^ After imaging, the digitally stored scan was manually analyzed by the sonographer using the calipers of the ultrasound scanner. SBP and DBP were measured at the supine position just before and immediately after the carotid ultrasound scanning using an automated Omron® M6 sphygmomanometer (Omron Matsusaka Co., Ltd, Kyoto, Japan). The mean of SBP and DBP values was used to calculate carotid elasticity. The diameter of the common carotid artery was measured at least twice both at end-diastole and at end-systole. The means of the measurements were used as the end-diastolic and end-systolic diameters. The ultrasound measurements and the concomitant brachial artery SBP and DBP measurements were used to calculate the following indices of carotid elasticity: cDIST=((Ds-Dd)/Dd)/(SBP-DBP), Young’s elastic modulus (YEM)=[(SBP−DBP)*Dd]/[(Ds−Dd)/IMT], and stiffness index (SI)= ln(SBP/DBP)/[(Ds−Dd)/Dd], where Dd is the diastolic diameter and Ds is the systolic diameter of the common carotid artery.^20^ All these indices of carotid elasticity describe the mechanical properties of the arterial wall. cDIST measures the ability of the carotid arteries to expand due to pressure changes induced by the cardiac cycle, YEM estimates the stiffness of the arterial wall, and SI describes the arterial wall stiffness independent of arterial BP.

### Assessment of childhood cardiometabolic risk factors

Whole body mass was measured twice, with the children having fasted for 12 h, emptied the bladder, and standing in light underwear by a calibrated InBody 720*^®^* bioelectrical impedance device (Biospace, Seoul, South Korea) to an accuracy of 0.1 kg. The mean of these two values was used in the analyses. Stature was measured three times with the children standing in the Frankfurt plane, without shoes, using a wall-mounted stadiometer to an accuracy of 0.1 cm. The mean of the nearest two values was used in the analyses. Body mass index was calculated by dividing body mass (kilograms) by body height (meters squared). BMI–SDS was calculated based on Finnish reference data.^21^ Waist circumference (WC) was measured three times after expiration at mid-distance between the bottom of the rib cage and the top of the iliac crest with an unstretchable measuring tape to an accuracy of 0.1 cm.^22^ Body fat percent (BF%) were measured by the Lunar*^®^* dual-energy X-ray absorptiometry device (GE Medical Systems, Madison, WI, USA) using standardized protocols.^23^

A research nurse took venous blood samples using a standard protocol the children having fasted for 12-h. Blood was immediately centrifuged, and plasma and serum samples were stored at a temperature of −75°C until biochemical analyses. Fasting plasma glucose concentration was measured using the hexokinase method (Roche Diagnostics GmbH, Mannheim, Germany), in which glucose is phosphorylated by hexokinase and the resulting NADPH formation is quantified spectrophotometrically at 340 nm. Fasting serum insulin concentration was measured by the electrochemiluminescence immunoassay with the sandwich principle (Roche Diagnostics GmbH, Mannheim, Germany). We calculated the HOMA-IR using the formula fasting plasma glucose (mmol/L) × fasting serum insulin (μlU/ml)/22.5.^24^

Fasting plasma concentrations of total cholesterol (TC), low density lipoprotein cholesterol (LDL-C), high density lipoprotein cholesterol (HDL-C), and triglycerides (TGs) were analyzed using a clinical chemistry analyzer (Hitachi High Technology Co, Tokyo, Japan). Large very low density lipoprotein cholesterol (large VLDL-C) was separated by ultracentrifugation (37 000 rpm, 15 h). Plasma concentrations of TC, LDL-C, HDL-C, large VLDL-C and TGs were measured by colorimetric enzymatic assays.^25^

The research nurse measured SBP and DBP manually from the right arm by a calibrated Heine 130 Gamma G7^®^ aneroid sphygmomanometer (Heine Optotechnik, Herrsching, Germany). The measurement protocol included a 5-min rest and thereafter three measurements in a sitting position at 2-min intervals. The mean of all three values was used as the SBP and DBP.^26^ The average BP was calculated by summing SBP and DBP and dividing the sum by two.

### Assessment of childhood biomarkers of inflammation and adipokines

Plasma high-sensitivity C-reactive protein (hsCRP) was measured using an enhanced immunoturbidimetric assay with the hsCRP (Latex) High Sensitivity Assay reagent (Roche Diagnostics GmbH, Mannheim, Germany) and the limit of quantitation of 0.3 mg/L. Plasma glycoprotein acetyls (GlycA) were analyzed using a high-throughput nuclear magnetic resonance (NMR) metabolomics platform (Nightingale Health Ltd, Kuopio, Finland).^27–29^ Plasma leptin was measured by a competitive radioimmunoassay (Multigamma 1261–001, PerkinElmer Wallac, Turku, Finland). Commercially available enzyme-linked immunosorbent assay (ELISA) kits were employed for the measurement of plasma interleukin-6 (IL-6) and tumor necrosis factor-alpha (TNF-α) (Sanquin Reagents Amsterdam, The Netherlands)^30^ and leptin receptor (BioVendor LM a.s., Brno, Chech Republic). Serum high molecular weight adiponectin (HMW-adiponectin) was analyzed using an ELISA kit after specific proteolytic digestion of other multimeric adiponectin forms (Millipore, Billerica, MA, USA).^30^

### Computation of childhood metabolic syndrome score and inflammation score

We computed a composite score called a metabolic syndrome score (MetSscore) by summing sex- and age-specific z-scores for the core components of MetS, including central adiposity (WC), IR (HOMA-IR), dyslipidemia (TGs and HDL-C combined), and elevated BP (SBP and DBP combined),^5,31^ using a formula zWC + zHOMA-IR + (zTG–zHDL)/2 + (zSBP+zDBP)/2.^5^ We used the average of the z-scores for SBP and DBP instead of the z-score for the mean arterial pressure (MAP) based on our previous statistical analyses on MetS,^32^ instead of the z-score for the mean arterial pressure (MAP) and the mean of the z-scores for TG and HDL-C, multiplying the z-score for HDL-C by −1 due to its inverse association with cardiometabolic risk, instead of the separate z-scores for TG and HDL-C. This approach was used to ensure that all core components of MetS are weighted equally in the MetSscore. A higher MetSscore reflects a less favorable cardiometabolic profile.

We computed a composite score for systemic low-grade inflammation summing biomarkers considered to reflect a pro-inflammatory state (hsCRP, GlycA, leptin, IL-6, TNF-α) and subtracting biomarkers considered to reflect an anti-inflammatory state (leptin receptor, HMW-adiponectin).33 We computed a modified inflammation score using population-specific *z*-scores for hsCRP, GlycA, leptin, IL-6, and TNF-α and a formula (zhsCRP + zGlycA + zleptin + zIL-6 + zTNF-α) - zleptin receptor - zHMW-adiponectin.^30^

### Statistical methods

All statistical analyzes were performed using the IBM SPSS statistics software, Version 28 (IBM Corp. NY, Armonk, USA). Before the analysis, the missing data on pubertal status at 8-year follow-up were replaced using multiple imputations, using 10 imputed data sets. Normality of variable distributions was evaluated visually from histograms and assessed using the Kolmogorov–Smirnov test. Differences in characteristics between girls and boys were assessed using the independent samples t-test. The associations of single cardiometabolic risk factors and the MetSscore at baseline with PWV, CAVI, cIMT, cDIST, YEM, and SI at the 8-year follow-up were analyzed using linear regression models adjusted for age and sex at baseline. The data were further adjusted for study group (intervention vs. control) and pubertal status at 8-year follow-up. We also studied if sex modified these associations by including sex x each cardiometabolic risk factor or the MetSscore interaction term into the linear regression models. The data from the linear regression analyses are presented as unstandardized regression coefficients (B), their 95% confidence intervals (CIs), and standardized regression coefficients (β). Analyses for PWV and CAVI were restricted to participants with complete data (n=175) because of missing data; given the limited number of covariates in the planned models, effect estimates are presented with 95% confidence intervals to describe their precision. Differences and associations with P-values <0.05 were considered statistically significant. The data were corrected for multiple testing using the Benjamini–Hochberg false discovery rate (FDR) with an FDR value of 0.2 (FDR^0.2^). We considered FDR corrected p-values ≤0.2 significant, as more stringent threshold would impair the balance between type I and type II errors.^34^ We estimated that 189 observations were needed to observe the beta estimate of 0.2 at the power of 0.80 when statistical significance level was set at p<0.05 in a three-predictor model.

## Results

### Participants

A total of 222 children (105 girls, 117 boys) aged 7-9 years had valid data on all required variables and were included in the final analyses. Data for the analyses on the associations of individual cardiometabolic risk factors and MetSscore at baseline with cIMT, cDIST, YEM, and SI at 8-year follow-up were available for 105 girls and 117 boys and on the associations of individual cardiometabolic risk factors and the MetSscore at baseline with PWV and CAVI at the 8-year follow-up were available for 85 girls and 90 boys. The baseline characteristics of the participants are presented in Table 1.

**Table 1.**
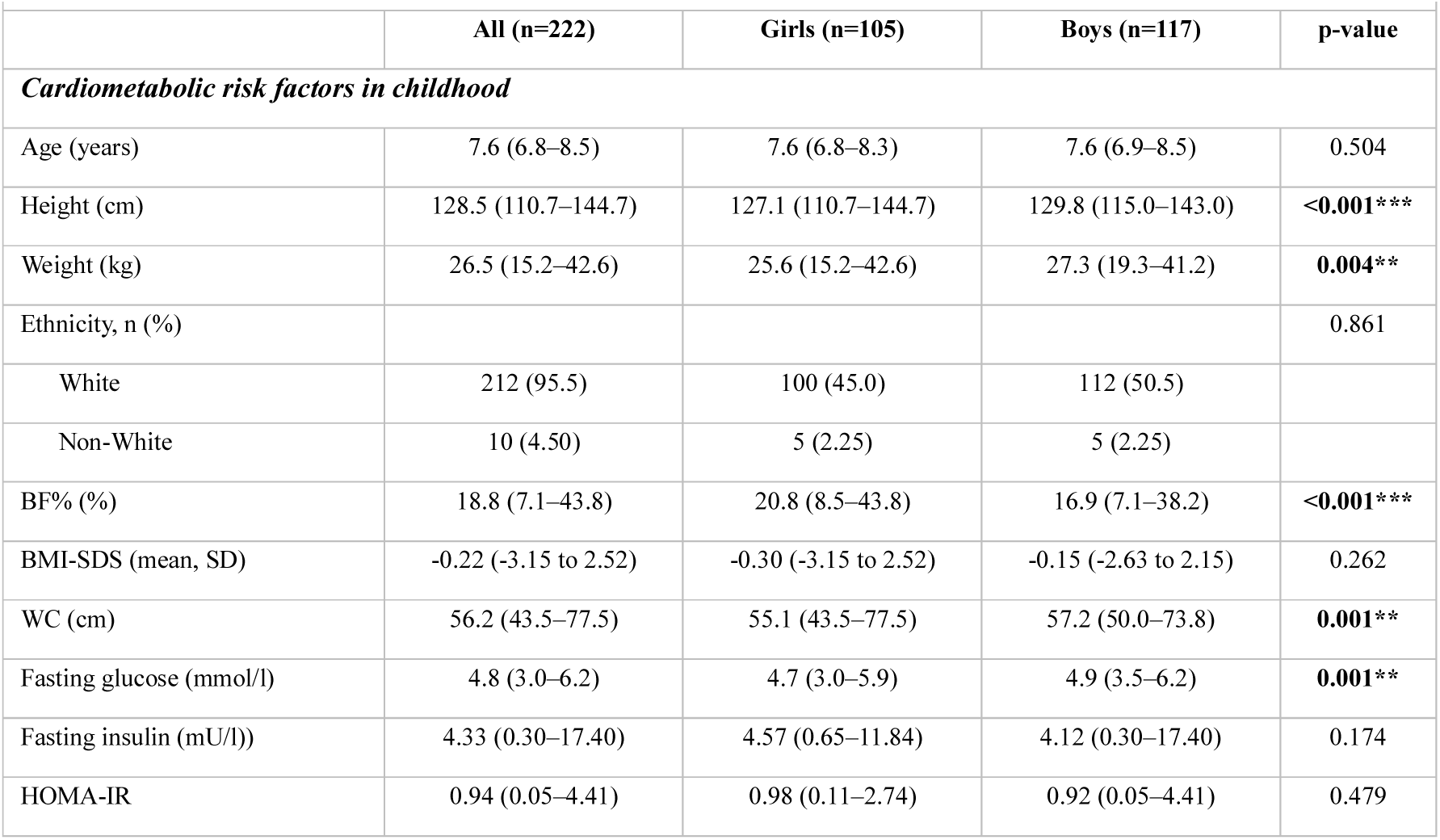
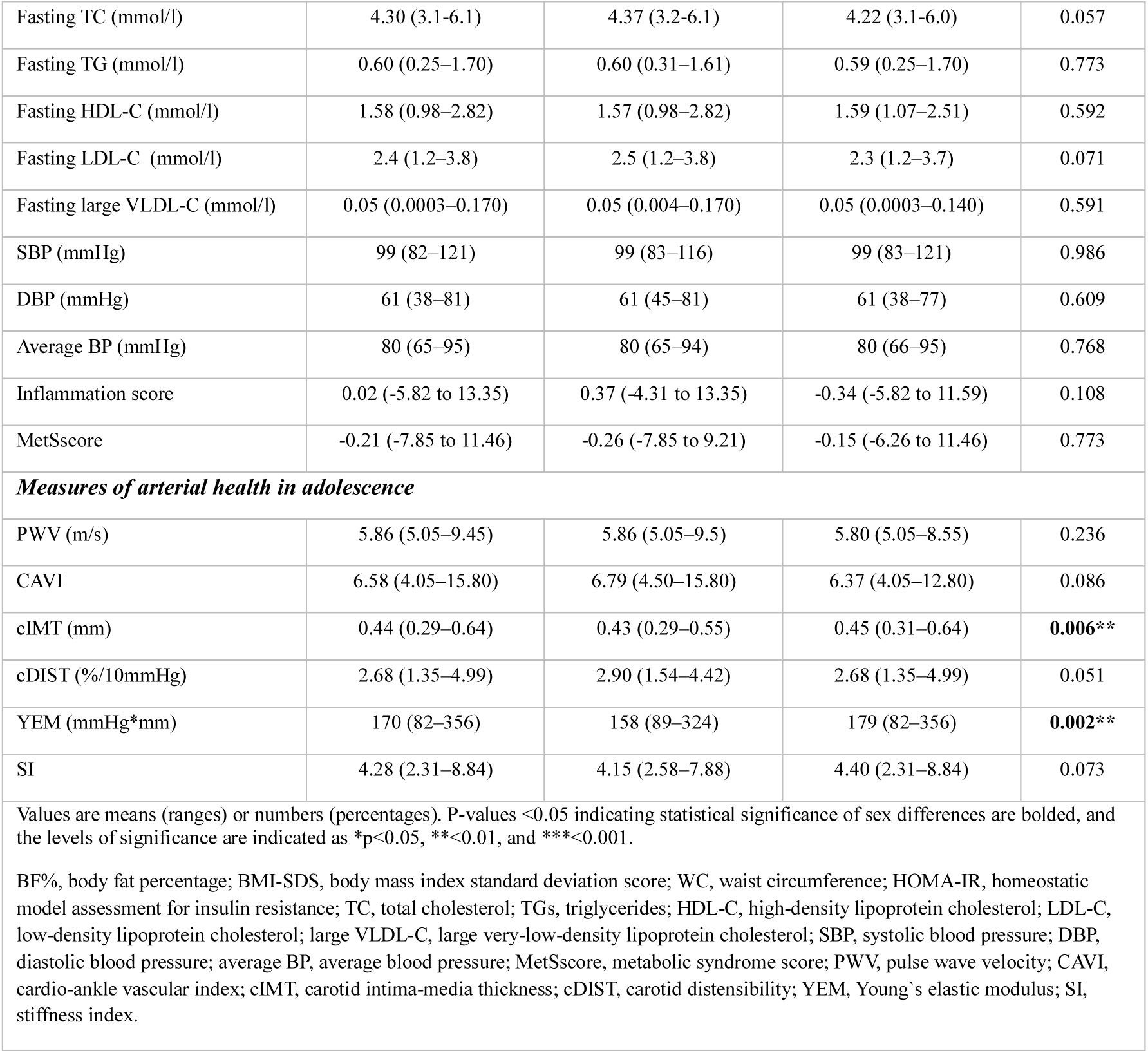
Baseline characteristics of the participants.

| Table 1. Baseline characteristics of the participants |  |  |  |  |
| --- | --- | --- | --- | --- |
|  | All (n=222) | Girls (n=105) | Boys (n=117) | p-value |
| *Cardiometabolic risk factors in childhood* |  |  |  |  |
| Age (years) | 7.6 (6.8–8.5) | 7.6 (6.8–8.3) | 7.6 (6.9–8.5) | 0.504 |
| Height (cm) | 128.5 (110.7–144.7) | 127.1 (110.7–144.7) | 129.8 (115.0–143.0) | <0.001*** |
| Weight (kg) | 26.5 (15.2–42.6) | 25.6 (15.2–42.6) | 27.3 (19.3–41.2) | 0.004** |
| Ethnicity, n (%) |  |  |  | 0.861 |
| White | 212 (95.5) | 100 (45.0) | 112 (50.5) |  |
| Non-White | 10 (4.50) | 5 (2.25) | 5 (2.25) |  |
| BF% (%) | 18.8 (7.1–43.8) | 20.8 (8.5–43.8) | 16.9 (7.1–38.2) | <0.001*** |
| BMI-SDS (mean, SD) | -0.22 (-3.15 to 2.52) | -0.30 (-3.15 to 2.52) | -0.15 (-2.63 to 2.15) | 0.262 |
| WC (cm) | 56.2 (43.5–77.5) | 55.1 (43.5–77.5) | 57.2 (50.0–73.8) | 0.001** |
| Fasting glucose (mmol/l) | 4.8 (3.0–6.2) | 4.7 (3.0–5.9) | 4.9 (3.5–6.2) | 0.001** |
| Fasting insulin (mU/l)) | 4.33 (0.30–17.40) | 4.57 (0.65–11.84) | 4.12 (0.30–17.40) | 0.174 |
| HOMA-IR | 0.94 (0.05–4.41) | 0.98 (0.11–2.74) | 0.92 (0.05–4.41) | 0.479 |
| Fasting TC (mmol/l) | 4.30 (3.1–6.1) | 4.37 (3.2–6.1) | 4.22 (3.1–6.0) | 0.057 |
| Fasting TG (mmol/l) | 0.60 (0.25–1.70) | 0.60 (0.31–1.61) | 0.59 (0.25–1.70) | 0.773 |
| Fasting HDL-C (mmol/l) | 1.58 (0.98–2.82) | 1.57 (0.98–2.82) | 1.59 (1.07–2.51) | 0.592 |
| Fasting LDL-C (mmol/l) | 2.4 (1.2–3.8) | 2.5 (1.2–3.8) | 2.3 (1.2–3.7) | 0.071 |
| Fasting large VLDL-C (mmol/l) | 0.05 (0.0003–0.170) | 0.05 (0.004–0.170) | 0.05 (0.0003–0.140) | 0.591 |
| SBP (mmHg) | 99 (82–121) | 99 (83–116) | 99 (83–121) | 0.986 |
| DBP (mmHg) | 61 (38–81) | 61 (45–81) | 61 (38–77) | 0.609 |
| Average BP (mmHg) | 80 (65–95) | 80 (65–94) | 80 (66–95) | 0.768 |
| Inflammation score | 0.02 (-5.82 to 13.35) | 0.37 (-4.31 to 13.35) | -0.34 (-5.82 to 11.59) | 0.108 |
| MetScore | -0.21 (-7.85 to 11.46) | -0.26 (-7.85 to 9.21) | -0.15 (-6.26 to 11.46) | 0.773 |
| *Measures of arterial health in adolescence* |  |  |  |  |
| PWV (m/s) | 5.86 (5.05–9.45) | 5.86 (5.05–9.5) | 5.80 (5.05–8.55) | 0.236 |
| CAVI | 6.58 (4.05–15.80) | 6.79 (4.50–15.80) | 6.37 (4.05–12.80) | 0.086 |
| cIMT (mm) | 0.44 (0.29–0.64) | 0.43 (0.29–0.55) | 0.45 (0.31–0.64) | **0.006\*\*** |
| cDIST (%/10mmHg) | 2.68 (1.35–4.99) | 2.90 (1.54–4.42) | 2.68 (1.35–4.99) | 0.051 |
| YEM (mmHg*mm) | 170 (82–356) | 158 (89–324) | 179 (82–356) | **0.002\*\*** |
| SI | 4.28 (2.31–8.84) | 4.15 (2.58–7.88) | 4.40 (2.31–8.84) | 0.073 |
| Values are means (ranges) or numbers (percentages). P-values <0.05 indicating statistical significance of sex differences are bolded, and the levels of significance are indicated as *p<0.05, **<0.01, and ***<0.001.<br>BF%, body fat percentage; BMI-SDS, body mass index standard deviation score; WC, waist circumference; HOMA-IR, homeostatic model assessment for insulin resistance; TC, total cholesterol; TGs, triglycerides; HDL-C, high-density lipoprotein cholesterol; LDL-C, low-density lipoprotein cholesterol; large VLDL-C, large very-low-density lipoprotein cholesterol; SBP, systolic blood pressure; DBP, diastolic blood pressure; average BP, average blood pressure; MetScore, metabolic syndrome score; PWV, pulse wave velocity; CAVI, cardio-ankle vascular index; cIMT, carotid intima-media thickness; cDIST, carotid distensibility; YEM, Young's elastic modulus; SI, stiffness index. |  |  |  |  |

### Associations of individual cardiometabolic risk factors and MetSscore in childhood with PWV and CAVI in adolescence

BF%, BMI-SDS, WC, HOMA-IR, fasting insulin, TGs, large VLDL-C, and the MetSscore in childhood were positively associated with PWV and CAVI in adolescence adjusted for age and sex at baseline (Table 2). However, the association between large VLDL-C and CAVI was not statistically significant after further adjustment for study group (p=0.476). SBP, average BP, and the inflammation score in childhood were positively associated with PWV in adolescence. Further adjustment for BF% attenuated the association between SBP in childhood and PWV in adolescence (B=0.017, 95% CI=0.006-0.029, β=0.216, p=0.004) but had no notable effect on other associations. These associations also remained after further adjustment for pubertal status and for FDR0.2 correction.

**Table 2.** Association of cardiometabolic risk factors in childhood with pulse wave velocity and cardio-ankle vascular index in adolescence among 175 participants.

|  | PWV |  |  |  | CAVI |  |  |  |
| --- | --- | --- | --- | --- | --- | --- | --- | --- |
| | B | 95% CI | $\beta$ | p-value | B | 95% CI | $\beta$ | p-value |
| BF% | 0.016 | 0.004 to 0.029 | 0.207 | **0.008** | 0.039 | 0.004 to 0.074 | 0.171 | **0.031** |
| BMI-SDS (kg/m<sup>2</sup>) | 0.109 | 0.024 to 0.195 | 0.187 | **0.013** | 0.250 | 0.003 to 0.496 | 0.150 | **0.047** |
| WC (cm) | 0.028 | 0.009 to 0.048 | 0.215 | **0.005** | 0.069 | 0.013 to 0.126 | 0.184 | **0.016** |
| Fasting glucose (mmol/l) | 0.145 | -0.088 to 0.378 | 0.096 | 0.221 | 0.291 | -0.378 to 0.960 | 0.068 | 0.392 |
| Fasting insulin (mU/l) | 0.064 | 0.026 to 0.101 | 0.246 | **0.001** | 0.161 | 0.053 to 0.270 | 0.217 | **0.004** |
| HOMA-IR | 0.270 | 0.190 to 0.430 | 0.242 | **0.001** | 0.685 | 0.221 to 1.150 | 0.216 | **0.004** |
| TC (mmol/l) | 0.012 | -0.136 to 0.161 | 0.012 | 0.872 | -0.089 | -0.514 to 0.337 | -0.031 | 0.681 |
| TG (mmol/l) | 0.545 | 0.196 to 0.894 | 0.229 | **0.002** | 1.477 | 0.473 to 2.481 | 0.218 | **0.004** |
| HDL-C (mmol/l) | 0.063 | -0.242 to 0.367 | 0.031 | 0.685 | -0.075 | -0.948 to 0.798 | -0.013 | 0.866 |
| LDL-C (mmol/l) | -0.033 | -0.196 to 0.131 | -0.030 | 0.693 | -0.119 | -0.588 to 0.349 | -0.038 | 0.616 |
| large VLDL-C (mmol/l) | 3.754 | 0.758 to 6.749 | 0.185 | **0.014** | 10.553 | 1.961 to 19.144 | 0.183 | **0.016** |
| SBP (mmHg) | 0.021 | 0.009 to 0.032 | 0.257 | **<0.001** | 0.034 | 0.000 to 0.068 | 0.147 | 0.053 |
| DBP (mmHg) | 0.005 | -0.007 to 0.017 | 0.065 | 0.391 | -0.003 | -0.037 to 0.030 | -0.014 | 0.852 |
| Average BP (mmHg) | 0.018 | 0.004 to 0.031 | 0.188 | **0.012** | 0.021 | -0.019 to 0.061 | 0.077 | 0.312 |
| Inflammation score | 0.034 | 0.007 to 0.060 | 0.188 | **0.013** | 0.53 | -0.024 to 0.129 | 0.103 | 0.177 |
| MetScore | 0.053 | 0.024 to 0.081 | 0.266 | **<0.001** | 0.124 | 0.041 to 0.207 | 0.219 | **0.004** |

Values are unstandardized regression coefficients B, their 95% confidence intervals, and standardized regression coefficients $\beta$ from linear regression models adjusted for age and sex.
**P-values <0.05** indicating statistical significance of associations **are bolded**.
BF%, body fat percentage; BMI-SDS, body mass index standard deviation score; WC, waist circumference; HOMA-IR, homeostatic model assessment for insulin resistance; TC, total cholesterol; TG, triglycerides; HDL-C, high density lipoprotein cholesterol; LDL-C, low density lipoprotein cholesterol; large VLDL-C, large very low density lipoprotein cholesterol; SBP, systolic blood pressure; DBP, diastolic blood pressure; average BP= average blood pressure; MetScore, metabolic syndrome score; PWV, pulse wave velocity; CAVI, cardio-ankle vascular index; cIMT, carotid intima media thickness; cDIST, carotid distensibility; YEM, Young's elastic modulus; SI, stiffness index

### Associations of individual cardiometabolic risk factors and MetSscore in childhood with cIMT, cDIST, YEM, and SI in adolescence

BF% and WC in childhood were positively associated with cIMT in adolescence adjusted for age and sex at baseline (Table 3). However, the association did not remain after the FDR0.2 correction (both corrected p-value 0.376). However, the association between WC and cIMT was not statistically significant after further adjustment for study group (p=0.060). HOMA-IR, fasting insulin, SBP, and average BP in childhood were inversely associated with cDIST in adolescence. BF%, BMI-SDS, WC, SBP, average BP and the MetSscore in childhood were positively associated with YEM in adolescence. The association between SBP in childhood and cDIST in adolescence (B=-0.015, 95% CI=-0.028 to -0.002, β=-0.154, p=0.024) and the association between HOMA-IR and cDIST (B=-0.180, 95% CI=-0.357 to -0.003, β=-0.145, p=0.046) weakened after further adjustment for BF%. The association between fasting insulin in childhood and cDIST in adolescence (B=-0.044, 95% CI=-0.087 to -0.001, β=-0.147, p=0.044) strengthened after further adjustment for BF%. The associations of single biomarkers of inflammation with measures of arterial health are presented in supplementary material. These associations also remained after further adjustment for pubertal status and for FDR0.2 correction.

**Table 3.**
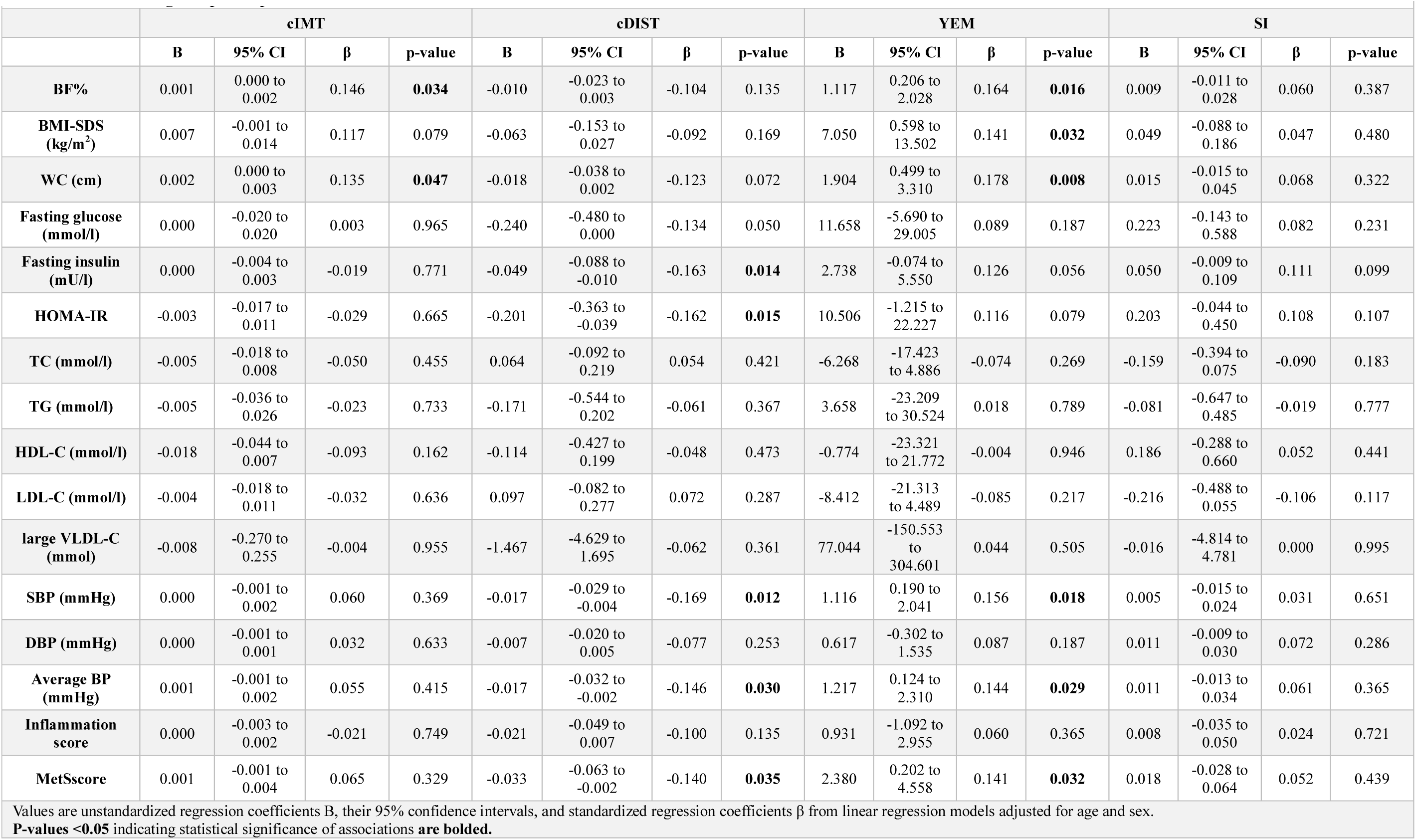
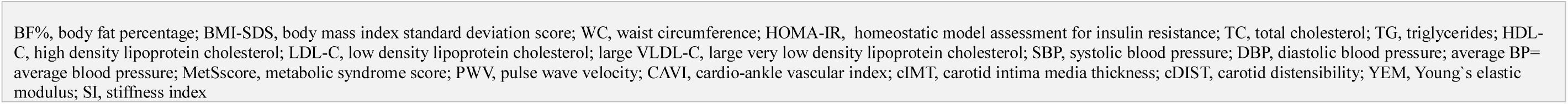
Association of cardiometabolic risk factors in childhood with risk factors with carotid intima-media thickness, carotid distensibility, Young’s elastic modulus and stiffness index in adolescence among 222 participants.

### Sex as a modifier for the associations of cardiometabolic risk factors in childhood with measures of arterial health in adolescence

DBP in childhood was positively associated with cIMT in adolescence in girls (B=0.002, 95% CI=0.001–0.003, β=0.293, p=0.003) but not in boys (B=-0.001, 95% CI=-0.003 to 0.000, β=0.166, p=0.074) adjusted for age at baseline (p=0.001 for interaction). Similarly, average BP in childhood was positively associated with cIMT in adolescence in girls (B=0.002, 95% CI=0.000–0.004, β=0.249, p=0.012) but not in boys (B=-0.001, 95% CI=-0.003 to 0.001, β=-0.101, p=0.279) adjusted for baseline age (p=0.016 for interaction).

## Discussion

We observed that childhood overall and abdominal adiposity, measured by BF%, BMI-SDS, and WC, associated with higher arterial stiffness, measured by PWV, CAVI, and YEM, eight years later in adolescence. Childhood BF% also associated with higher cIMT in adolescence. These findings support previous evidence that excess adipose tissue association of higher arterial stiffness in children and adolescents^35,36^ and that childhood overweight is associated with increased cIMT in adulthood.^37^ Circulating biomarkers of systemic low-grade inflammation hsCRP and GlycA as well as circulating adipokines leptin and adiponectin have been positively associated with arterial stiffness^38^ and endothelial dysfunction in youth.^39^ Partly supporting these findings, we observed that the inflammation score and leptin in childhood were positively and childhood leptin receptor was inversely associated with PWV, CAVI, YEM, and SI but none of the single biomarkers of inflammation in childhood was associated with cIMT in adolescence. Moreover, leptin receptor was positively associated with cDIST. These findings suggest that inflammation may increase arterial stiffness in youth but that a longer-term exposure to inflammation until adulthood may be required for structural atherosclerotic changes in artery walls.

Previous studies have shown a strong association between IR and higher arterial stiffness among children and adolescence.^40,41^ In line with the results of these studies,^40,41^ we also found that IR in childhood associated with higher PWV and CAVI and lower cDIST, indicating higher arterial stiffness and poorer arterial elasticity, in adolescence. One explanation for these observations could be that IR initiates a cascade from hyperinsulinemia to endothelial dysfunction, oxidative stress, systemic low-grade inflammation and finally arterial stiffening.^42^ IR has also been positively associated with circulating leptin, TNF-α, and IL-6 and inversely with circulating adiponectin, suggesting that IR results in a proinflammatory state and impairs arterial function in adults.^43,44^

VLDL transports triglycerides from the liver to adipose tissue and muscles, LDL delivers cholesterol from the liver to peripheral tissues, and HDL removes excess cholesterol from peripheral tissues and delivers it to the liver for excretion, these lipoprotein particles thus collectively contributing to lipid metabolism. We observed that mild childhood dyslipidemia, indicated by elevated plasma levels of TGs and VLDL-C, associated with higher PWV and CAVI in adolescence. This finding suggests that dyslipidemia plays an important role in early arterial dysfunction. Consistent with our results, plasma levels of VLDL-C and TG-rich VLDL particles were positively associated with arterial stiffness in a cross-sectional study among 10-24 year-olds.^45^ Moreover, childhood adiposity has been observed to associate not only increased plasma LDL-C and decreased plasma HDL-C but also increased carotid artery stiffness measured by cIMT, YEM and SI in adulthood.^37,46^ Increased plasma LDL-C and decreased plasma HDL-C were also found to association of increased cIMT from adolescence to early adulthood.^47^ However, we did not observe an association of plasma LDL-C or HDL-C in childhood with any measure of arterial structure or function in adolescence. Although increased LDL-C is a major risk factor for atherosclerosis in adulthood,^48^ the short exposure of artery walls to the relatively low plasma LDL-C levels in youth is likely to explain the absence of an association between plasma LDL-C and any arterial measure in our longitudinal study lasting from childhood to adolescence. Moreover, the ability of increased HDL particles to decrease systemic low-grade inflammation and protect against atherosclerosis in any age group is likely to be determined primarily by its structure and composition rather than by its plasma levels.^49^ Large, TG-rich VLDL particles are highly pro-inflammatory and can impair arterial endothelial function, and unlike LDL particles, are able to cross the endothelium with unoxidized particles.^50^ Together with this evidence, our findings suggest that higher TGs and large VLDL-C are better predictors for early signs of atherosclerosis than higher LDL-C or lower HDL-C in youth.^51^

Previous studies have shown that BP, particularly central SBP, is positively associated with PWV, cIMT and inversely associated with cDIST in children and adolescents.^52,53^ Consistent with these findings, we observed that childhood SBP and average BP were positively associated with PWV in adolescence. Central SBP has also been positively associated with cIMT in previous studies among children and adolescents with type 1 diabetes,^53^ whereas we did not find an association of SBP or average BP in childhood with cIMT in adolescence. One explanation for this discrepancy may be that PWV and CAVI are more sensitive indicators of early arteriosclerotic changes than cIMT, a measure of arterial wall thickness, and cDIST and YEM, measures of arterial elasticity.^54^ It should also be noted that PWV and CAVI measure arterial stiffness and elasticity throughout the arterial tree, especially in aorta and other large arteries, whereas cIMT reflects local thickening in the wall of the common carotid artery,^11,16^ that may partly explain why BP was related to arterial stiffness but not carotid wall thickness in our study.

We found that higher DBP and average BP in childhood were associated with higher cIMT in adolescence among girls but not in boys. This sex difference may reflect greater sensitivity of girls to the effects of BP on artery walls due to their higher circulating levels of sex hormones,^55^ a more advanced puberty,^55^ or other biological differences^56^ in late childhood and adolescence compared to boys. Estrogens and testosterone influence arterial structure, fat distribution, and immune responses during sexual maturation, and these effects may differ between sexes.^57^ cIMT increases in children and adolescents with age, pubertal development, and growth,^58,59^ and this change is influenced by sex.^49,55,58^ cIMT varies across pubertal stages in childhood and adolescence being lowest in prepuberty and highest after puberty.^55^ Moreover, earlier onset of puberty has been associated with higher BP and increase the risk of developing ASCVDs in adults^60^ The American Heart Association recommends ambulatory BP monitoring in children and adolescents at increased risk of hypertension and ASCVDs, as it better reflects real-world situations and is more accurate in assessing true BP levels than clinic BP measurement.^61,62^ Moreover, regular BP monitoring is important in children and adolescents at increased risk of ASCVDs, since elevated BP values may predispose to early structural and functional changes in the arteries and predict future risk. It should be noted, however, that a slightly increased arterial IMT in youth is mainly due to thickening of the media layer and may reflect normal arterial remodeling in response to increased arterial pressure and flow but not early atherosclerosis.^63,64^

Our research group has previously found a positive cross-sectional association between the MetSscore and arterial stiffness in 6-8-year-old children.^4^ However, the results of longitudinal studies in adults suggest that a prevalent MetS or MetSscores do not consistently outperform single cardiometabolic risk factors in predicting the development of atherosclerosis or ASCVDs^65,66^ In the current longitudinal study, we observed slightly stronger positive associations of the MetSscore than most of the single cardiometabolic risk factors in childhood with PWV, CAVI, and YEM in adolescence. Therefore, our findings indicate that assessing the MetSscore in childhood could help identify children with not only MetS but also early signs of atherosclerosis and thereby target and tailor lifestyle interventions to those who benefit most to prevent the development of ASCVDs in later life.

The strengths of our study include the population-based sample of children followed up for eight years until adolescence and the availability of several measures of cardiometabolic and arterial health enabling a comprehensive analysis of predictors for early signs of atherosclerosis in youth. A limitation of our study is that the participants were mainly White, which limits the generalizability of our findings to other pediatric populations. Finally, we had comprehensive data on arterial structure and function only in adolescence, which did not allow us to study the associations of changes in cardiometabolic risk factors with changes in arterial health from childhood to adolescence.

In conclusion, our study suggest that greater body adiposity, IR, higher BP, and clustering of cardiometabolic risk factors in childhood associated with arterial stiffness and reduced arterial distensibility in adolescence. These findings highlight the importance of identification of children and adolescents with MetS or single cardiometabolic risk factors and thereby target and tailor lifestyle interventions to those who benefit most to prevent the development of ASCVDs in later life. Moreover, our results support the development and implementation of effective preventive strategies and health policies to reduce cardiometabolic risk and long-term disease burden. Future research is needed to explore how and which lifestyle changes in youth affect arterial health later in life.

## Non-standard Abbreviations and Acronyms

ASCVD: atherosclerotic cardiovascular disease
BF%: body fat percentage
BMI-SDS: body mass index standard deviation score
BP: blood pressure
CAVI: cardio-ankle vascular index
CI: confidence interval
cDIST: carotid distensibility
cIMT: carotid intima-media thickness
DBP: diastolic blood pressure
ELISA: enzyme-linked immunosorbent assay
FDR: false discovery rate
GlycA: glycoprotein acetyls
HDL-C: high-density lipoprotein cholesterol
HMW-adiponectin: high molecular weight adiponectin
HOMA-IR: homeostatic model assessment for insulin resistance
hsCRP: high-sensitivity C-reactive protein
IL-6: interleukin-6
IR: insulin resistance
LDL-C: low-density lipoprotein cholesterol
MAP: mean arterial pressure
MetS: metabolic syndrome
MetSscore: metabolic syndrome score
NMR: nuclear magnetic resonance
PANIC: Physical Activity and Nutrition in Children
PWV: pulse wave velocity
SBP: systolic blood pressure
SI: stiffness index
TC: total cholesterol
TG / TGs: triglyceride(s)
TNF-α: tumor necrosis factor alpha
VLDL-C: very-low-density lipoprotein cholesterol
WC: waist circumference
YEM: Young’s elastic modulus

## Acknowledgements

The PANIC study has been supported by grants from the Research Council of Finland, Ministry of Education and Culture of Finland, Ministry of Social Affairs and Health of Finland, Research Committee of the Kuopio University Hospital Catchment Area (State Research Funding), Finnish Innovation Fund Sitra, Social Insurance Institution of Finland, Finnish Cultural Foundation, Foundation for Paediatric Research, Diabetes Research Foundation in Finland, Finnish Foundation for Cardiovascular Research, Juho Vainio Foundation, Paavo Nurmi Foundation, Yrjö Jahnsson Foundation, and the city of Kuopio.

## Sources of Funding

The PANIC study has been supported by grants from the Research Council of Finland, Ministry of Education and Culture of Finland, Ministry of Social Affairs and Health of Finland, Research Committee of the Kuopio University Hospital Catchment Area (State Research Funding), Finnish Innovation Fund Sitra, Social Insurance Institution of Finland, Finnish Cultural Foundation, Foundation for Pediatric Research, Diabetes Research Foundation in Finland, Finnish Foundation for Cardiovascular Research, Juho Vainio Foundation, Paavo Nurmi Foundation, Yrjö Jahnsson Foundation, and the city of Kuopio.

## Disclosures

None declared.

## Data Avaibility Statement

Information about the PANIC study and the data used in the present paper is available at www.panicstudy.fi/en/etusivu. The data are not publicly available due to research ethical 21 reasons and because the owner of the data is the University of Eastern Finland and not the research group. However, the corresponding author can provide further information on the PANIC study and the PANIC data on a reasonable request.

## Notes

### Competing Interest Statement

The authors have declared no competing interest.

### Funding Statement

This research received no external funding. The authors and their institution did not receive any payments, support, or services from third parties for any aspect of the submitted work.

